# Induction of an early IFN-γ cellular response and high plasma levels of SDF-1α are inversely associated with COVID-19 severity and residence in rural areas in Kenyan patients

**DOI:** 10.1101/2024.12.22.24319505

**Authors:** Perpetual Wanjiku, Benedict Orindi, John Kimotho, Shahin Sayed, Reena Shah, Mansoor Saleh, Jedidah Mwacharo, Christopher Maronga, Vivianne Olouch, Ann Karanu, Jasmit Shah, Zaitun Nneka, Lynette Isabella Ochola-Oyier, Abdirahman I. Abdi, Susanna Dunachie, Philip Bejon, Eunice Nduati, Francis M. Ndungu

## Abstract

**Introduction:** COVID-19 was less severe in Sub-Saharan Africa (SSA) compared with Europe and North America. It is unclear whether these differences could be explained immunologically. Here we determined the levels of *ex vivo* SARS-CoV-2 peptide-specific IFN-γ producing cells, and levels of plasma cytokines and chemokines over the first month of COVID-19 diagnosis among Kenyan COVID-19 patients from urban and rural areas.

**Methods:** Between June 2020 and August 2022, we recruited and longitudinally monitored 188 COVID-19 patients from two regions in Kenya, Nairobi (urban, n = 152) and Kilifi (rural, n = 36), with varying levels of disease severity – severe, mild/moderate, and asymptomatic. IFN-γ secreting cells were enumerated at 0-, 7-, 14- and 28-days post diagnosis by an *ex vivo* enzyme-linked immunospot (ELISpot) assay following *in vitro* stimulation of peripheral blood mononuclear cells (PBMCs) with overlapping peptides from several SARS-CoV-2 proteins. A multiplexed binding assay was used to measure the levels of 22 plasma cytokines and chemokines.

**Results:** Higher frequencies of IFN-γ-secreting cells against the SARS-CoV-2 spike peptides were observed on the day of diagnosis among the asymptomatic compared to the patients with severe COVID-19. Higher concentrations of 17 of the 22 cytokines and chemokines measured were positively associated with severe disease, and in particular interleukin (IL)-8, IL-18 and IL-1ra (p<0.0001), while a lower concentration of SDF-1α was associated with severe disease (p<0.0001). Concentrations of 8 and 16 cytokines and chemokines including IL-18 were higher among Nairobi asymptomatic and mild patients compared to their respective Kilifi counterparts. Conversely, the concentrations for SDF-1α were higher in rural Kilifi compared to Nairobi (p=0.012).

**Conclusion:** In Kenya, as seen elsewhere, pro-inflammatory cytokines and chemokines were associated with severe COVID-19, while an early IFN-γ cellular response to overlapping SARS-CoV-2 spike peptides was associated with reduced risk of disease. Living in urban Nairobi (compared with rural Kilifi) was associated with increased levels of pro-inflammatory cytokines/chemokines.

## Introduction

Despite widespread transmission of SARS-CoV 2, SSA experienced a reduced burden of severe coronavirus disease 2019 (COVID-19) and associated mortality than North America and Europe (1). This observation is both puzzling (2) and paradoxical (3), because of the relatively weaker and underfunded health systems in SSA. Some of the proposed, but still unproven, explanations for the reduced burden of severe COVID-19 in SSA include under-diagnosis of cases and mortality, a younger population (7), warmer climatic conditions with outdoor living, high levels of pre-existing cross-protective antibodies and T-cells induced from a high prevalence of infectious agents with SARS-CoV-2 like immune determinants, and immune regulation associated with either prior BCG vaccination (5–11) or chronic/repeated infections including helminths (12) and malaria (13). Like other viruses, SARS-CoV-2 induces a plethora of inflammatory host responses, including cytokines and chemokines (14), that play key roles in either protective immunity or immunopathology (15). Notably, the pathogenesis of COVID-19 has been linked to dysregulated and excessive cytokine and chemokine responses, upon SARS-CoV-2 infection (16). Numerous studies have linked increased levels of cytokines and chemokines to severe COVID-19 and associated mortality, including IL-1β, IL-1ra, IL-2, IL-6, IL7, IL-8, IL-18, IFN-γ, TNF-α, IFN-γ-inducible protein 10 (IP-10), granulocyte macrophage-colony stimulating factor (GM-CSF), monocyte chemoattractant protein-1 (MCP-1), and Macrophage inflammatory protein-1 alpha (MIP-1-α) (17–24). Collectively called a “cytokine storm”, a dysregulated cytokine response is implicated as the cause of the multiple organ failures and the acute respiratory distress syndrome (ARDS), which characterise severed COVID-19 and associated fatalities (25).

Published data show that the characteristic antibody response to SARS-CoV-2 infection, where levels increase with time, viral loads and COVID-19 severity, were experienced in Kenyan patients (26). Thus, there is no evidence of the primary acute antibody response controlling the infection outcome. Aside from antibodies, T-cell responses can control viremia, either directly by killing virus infected cells or indirectly by providing the relevant co-stimulatory molecules for supporting antibody production by B cells (27). However, there is a paucity of data on the cellular response to SARS-CoV-2 in Kenyan patients, and its possible role in modulating disease severity. Furthermore, it is unclear whether the differences in the rates of severe disease between urban and rural dwellers within SSA, as well as between developed countries and SSA, could be explained immunologically. In this study, we collected longitudinal blood samples from individuals from Nairobi (urban) and Kilifi (rural) with varying degrees of COVID-19 severity (asymptomatic, mild/moderate and severe) and compared levels of their *ex vivo* SARS-CoV-2 Spike peptide-specific IFN-γ producing cells, and levels of plasma cytokines and chemokines over their first month of COVID-19 diagnosis.

## Methods

### Study design, setting and participants

Participant sampling has been described previously (26). Briefly, we included 400 blood samples from a longitudinal cohort study of 188 patients that aimed at understanding the kinetics of naturally acquired immune responses to SARS-CoV-2 among COVID-19 patients from two sites in Kenya: 1) The Aga Khan University Hospital (AKUH) in Nairobi, an urban metropolitan academic medical Centre; and 2) Kilifi County Hospital, a community-based government hospital serving a rural coastal region. The samples were collected during the COVID-19 pandemic between June 2020 and August 2022. At the time of the study, SARS-CoV-2 transmission in Nairobi was higher than in Kilifi (28). Participants were adults, aged ≥18 years old, recruited within seven days of positive diagnosis of COVID-19 by RT-PCR testing. Initial sampling was at day 0 (i.e., day of diagnosis). Follow-up and subsequent samplings were done on days 7, 14 and 28 from a positive SARS-CoV-2 infection diagnosis. We collected 20 mL of venous blood in sodium heparin vacutainer. Additionally, we included residual longitudinal plasma samples from the AKUH biobank for cytokine and chemokine measurements. These samples were collected from COVID-19 patients who consented to this follow-up study on the day of diagnosis (i.e., day 0) and on day 28.

### COVID-19 severity classification

We included patients from five COVID-19 severity groups of asymptomatic, mild, moderate, severe, and critical as determined by clinicians at the time of diagnosis following the National Institutes of Health (NIH, USA) guidelines(29). Asymptomatic patients were those who tested positive for SARS-CoV-2 via RT-PCR but did not display any COVID-19 symptoms. Mild cases were SARS-CoV-2 positive and exhibited symptoms such as fever, sore throat, cough, malaise, headache, muscle pain, vomiting, nausea, diarrhoea, anosmia, or ageusia, without any signs of shortness of breath, dyspnea, or abnormal chest imaging. Moderate cases tested positive for SARS-CoV-2 and showed evidence of lower respiratory tract infection based on clinical examination or imaging, with an oxygen saturation (SpO2) of ≥94% on room air. Severe cases were positive for SARS-CoV-2 with an SpO2 of <94% on room air, a ratio of arterial partial pressure of oxygen to fraction of inspired oxygen (PaO2/FiO2) <300 mm Hg, a respiratory rate >30 breaths/minute, and/or lung infiltrates >50%. Critically ill patients were positive for SARS-CoV-2 and experienced respiratory failure, septic shock, and/or multiple organ dysfunction syndrome. Due to the small numbers in the moderate and critical groups, mild cases were lumped together with moderate (mild/moderate), whilst critical were grouped with severe ones (severe). Thus, we studied immune responses among three COVID-19 patient groups: asymptomatic, mild/moderate and severe.

### Procedures

#### Plasma separation and PBMC isolation

Plasma was separated by centrifuging the tubes at 440 g for 10 minutes using an Eppendorf 5810R centrifuge, aliquoted in 2 mL microcentrifuge tubes, and immediately stored in -80°C freezers until the time for laboratory analysis. PBMCs were isolated from the remaining blood component using density gradient centrifugation media (Lymphoprep^TM^ (1.077 g/ml, Stem Cell Technologies), aliquoted to 1.8 mL cryovials, and stored in -196°C liquid nitrogen tanks until usage. Plasma and PBMC samples from AKUH were transported in dry ice and liquid nitrogen respectively, to the KEMRI–Wellcome Trust Research Programme laboratories in Kilifi and stored appropriately for laboratory analyses. Prior to use, PBMCs were thawed and rested at 37°C, 5% CO_2_ for 15–16 hours. PBMC counting was done using Vi-CELL XR Cell Viability Analyser or Countess™ Cell Counting Chamber Slides (Thermo Scientific) before assay setup.

#### SARS-CoV-2 synthetic peptides pools for ELISpot measurements

A total of 641 peptides (15–18-mers with a ten amino acid overlap) were pooled into ten peptide pools, spanning different regions of the virus. The pools covered: the spike protein region (S1: positions 1–93 and S2: positions 94–178), membrane protein (M: positions 1–31), nucleocapsid protein (NP: positions 1–55), non-structural proteins (NSP 3B: positions 207– 306, NSP 3C: position307–379, NSP 12B: positions 665–729 and NSP 15–16: positions 886–972) and Open Reading Frame (ORF3: positions 1–37 and ORF8: positions 1–15). These peptides (which were synthesised by Mimotopes Pty Ltd and a kind donation from Professor Susanna Dunachie’s laboratory, Oxford University) were used to stimulate PBMCs for *ex vivo* IFN-γ ELISpot assay. The peptide sequences and pooling details are provided in Table S1.

#### Interferon gamma ELISpot assay

To quantify *the ex vivo* interferon gamma (IFN-γ) cellular response to overlapping SARS-CoV-2 peptides from different proteins, we stimulated PBMC with synthetic peptides pools through an *in vitro* IFN-γ ELISpot assay, as previously described (30). Briefly, 5 μg/ml of anti-human IFN-γ antibody clone 1-D1K (Mabtech, AB, Sweden) was used to coat Multiscreen-I 96 ELISpot plates overnight. Individual’s PBMC were plated in duplicates at 200,000 cells per well for each specific protein, based on pre-prepared plate templates.

Peptide pools were then added at a final concentration of 2 µg/mL per wells and incubated for 16–18 hours at 37^0^C, 5% CO_2_, 95% humidity. Concanavalin A (ConA; Sigma) was used as the positive control at a final concentration of 5 µg/mL per well, while dimethyl sulfoxide (DMSO; Sigma), which was a constituent of the diluent for the peptides and Con A was used at a similar concentration to peptides to serve as the negative control. IFN-γ secreting cells were then detected using an anti-human IFN-γ biotinylated antibody clone 7-B6-1 (Mabtech) at 1 μg/mL and an incubation for 2–4 hours. Thereafter, streptavidin alkaline phosphatase antibody (Mabtech) was added at 1 μg/mL and incubated for 1–2 hours, and the IFN-γ spots then developed using 1-Step^TM^ NBT/BCI (nitro blue tetrazolium/5-bromo-4-chloro-3-phosphatase) substrate (Thermo Scientific) during a 7-minute incubation in the dark. The enzyme-substrate reaction was stopped by rinsing the plate 3 times under running tap water. Plates were then airdried for at least 2 days on an open lab bench, and spots enumerated on an AID ELISpot Reader version 4.0. Results are hereby reported as spot-forming units (SFU)/10^6 PBMC after subtracting the background (mean SFU from negative control wells). Data from failed individual PBMC tests, defined here as either, an excessive background where the negative control wells had >80 SFU/10^6 PBMCs, or a positive control well with an average of <100 SFU/10^6 PBMCs (too few), were excluded. We also applied the ELISpot assay limit of detection of 10 SFU/10^6 PBMCs, with all wells having values <10 SFU/10^6 PBMCs replaced with 5 SFU/10^6 PBMCs. For the participants who did not have enough PBMC to be tested against all the peptide pools, we prioritised measurements against pools from S1, S2, NP and M proteins. Data are reported only for the individuals whose PBMC were tested against all the available peptide pools for each specific protein segment. We summed the responses from the different pools of the same protein segment, which resulted in different sample sizes for different proteins as follows: spike (171 samples for S1, S2), NP (162 samples), M (136 samples), NSPs (100 samples for NSP 3B, NSP 3C, NSP 12B and NSP 15 -16) and ORF (90 samples for ORF 3 and ORF 8) Table S2.

#### Luminex assay

Plasma concentrations of 22 cytokines and chemokines were measured using a Human ProcartaPlex™ Human Panel 1A (Thermo Fisher Scientific, Cat. No. EPX010-12010-901, Lot number 316776-000), which consisted of: a) Th1/Th2 specific cytokines: GM-CSF, IFN-γ, IL-1β, IL-2, IL-6, IL-8, IL-18, TNF-α, IL-9, IL-21; b) Pro-Inflammatory cytokines: IFN-α, IL-1α, IL-1ra, IL-7, TNF-β; and c) Chemokines: Eotaxin, GRO-α, IP-10, MCP-1, MIP-1α, MIP-1β, SDF-1α, according to the manufacturer’s instructions. Briefly,1x capture magnetic beads were added to the plates, and unbound beads were then washed away with 1X wash buffer. Plasma samples were thawed and diluted at 1:2 with 1X universal assay buffer (UAB) before addition to the plates. Standards from the kit at 4-fold serial dilutions of 1:5, 1:20, 1:80, 1:320, 1:1280, 1:5120, 1:20480 as well as a blank (UAB), were also added. The plates were then incubated on a shaker at 600 rpm for 2 hours at room temperature. After incubation, contents were discarded, plates washed, and 1X biotinylated detection antibody added. The plates were then incubated for 30 minutes on an Eppendorf Thermomixer Comfort shaker at room temperature. The plates were then washed, and 1X Streptavidin-PE added and incubated for 30 minutes on an Eppendorf Thermomixer Comfort shaker at room temperature, the plates were then washed before adding 1X reading buffer for 5 minutes on an Eppendorf Thermomixer Comfort shaker at room temperature. All wash steps were performed on an Invitrogen hand-held magnetic plate washer. Data were acquired on the Magpix systems multiplex Luminex machine and concentrations (pg/mL) of the samples calculated in Belysa® Immunoassay Curve Fitting software version 1.1.0 (31) using a 5- or 4-parameter logistic standard curve generated from standards of known concentration.

### Statistical analysis

For the ELISpot data, time point specific geometric means (GMs) of IFN-γ secreting cells for each of the different SARS-COV-2 peptides pools were calculated for each severity group and geographical location. For each peptide, variations in ELISpot responses were compared using a linear mixed effects model on log-transformed PBMCs values with patient as a random effect, and time (i.e., day of sampling), severity group and time-by-severity group interaction term as fixed effects (32), followed by Tukey’s multiple comparisons. Within each severity group, differences between geographic locations were compared using Kruskal Wallis paired with a Dunn’s multiple comparisons. This analysis set was restricted to 59 patients who had PBMC samples.

Cytokine and chemokine data were first normalised to have a zero mean and a standard deviation of one, and cross-correlations among the cytokines determined using Pearson correlations and principal component analysis (PCA). Principal components (PCs) were extracted based on scree-plot, variance explained and the interpretability of the components. Next, the non-normalised cytokine and chemokine data were log-transformed and fitted into linear mixed effects regression models with age, sex, day of sampling, location or disease severity as fixed effects and patient as a random effect. Interactions were explored and separate models fitted where necessary. For location comparisons, severe and moderate COVID-19 cases were excluded as these categories were only present in the Nairobi data.

Results from the regression models were presented using heat maps, in which the effect size (i.e., coefficient) determined the density of colour-shading for each square. TNF-β data were excluded from the analyses as only one patient had a measurable concentration. Thus, we analysed 21 cytokines from all 188 patients. Analyses were performed using R version 4.3.0 (33). The factoextra package (34) was used for PCA. For visualisations, the pheatmap (35) and ggplot2 (36) packages and GraphPad Prism Software version 10.1.2 (37) were used.

### Ethical approval

The study obtained ethical approval from the Kenya Medical Research Institute’s Scientific and Ethics Review Unit (KEMRI SERU; protocol no. 4081) and the Aga Khan University, Nairobi, Institutional Ethics Review Committee (protocol no. 2020 IERC-135 V2). Written informed consent was obtained from all willing patients before their enrolment into the study.

## Results

### Participant baseline characteristics

Using the NIH clinical guidelines for grading COVID-19 severity of 188 patients, 27 (14%) were asymptomatic, 75 (40%) were mild/moderate and 86 (46%) were severe cases. Collectively, the 188 patients contributed 400 blood samples collected over the first month of diagnosis: day 0 (187 samples), day 7 (50 samples), day 14 (53 samples), and day 28 (110 samples). Fewer samples were collected on days 7 and 14 mainly due to design; that is, residual longitudinal plasma samples from AKUH biobank were only collected in day 0 and day 28. Of the 188 patients, 129 (69%) were male, 36 (19%) were from rural Kilifi and 152 (81%) from urban Nairobi (Table 1). Their median age at recruitment was 48 years (IQR 37– 58), with disease severity increasing with age. All 86 patients with severe COVID-19 were from Nairobi as we were unable to recruit severe patients in Kilifi. Underlying comorbidities such as diabetes, HIV/AIDS, and hypertension were prevalent among the mild/moderate and severe COVID-19 groups. Two participants with severe disease died on day 7 and 28 respectively (Table 1).

**Table 1.** Participant demographic and clinical characteristics^†^

| Characteristic | Asymptomatic<br>(n = 27) | Mild/Moderate<br>(n = 75) | Severe<br>(n = 86) |
| --- | --- | --- | --- |
| Median age at recruitment<br>(IQR), years | 40 (33, 55) | 46 (35, 54) | 53 (43, 60) |
| Male sex | 15 (56%) | 46 (61%) | 68 (79%) |
| Location |  |  |  |
| Kilifi | 21 (78%) | 15 (20%) | 0 |
| Nairobi | 6 (22%) | 60 (80%) | 86 (100%) |
| Comorbidities |  |  |  |
| Diabetic | 1 (7%) | 28 (42%) | 47 (63%) |
| Hypertensive | 0 | 15 (22%) | 33 (44%) |
| HIV positive | 0 | 5 (8%) | 4 (5%) |
| Had COPD | 0 | 0 | 1 (1%) |
| Had Renal disease | 0 | 0 | 2 (3%) |
| Had heart disease | 0 | 0 | 6 (8%) |
| Patient management |  |  |  |
| Were hospital admissions | 0 | 56 (84%) | 75 (100%) |
| In intensive care unit | 0 | 4 (6.0%) | 15 (20%) |
| Clinical Outcome, died | 0 | 0 | 2 (3%) |

† Data are median (IQR) or number (%); COPD if Chronic obstructive pulmonary disease.

### Kinetics and frequencies of *ex vivo* IFN-γ secreting cells by COVID-19 severity and geographical location

Kinetics and levels of IFN-γ secreting cell responses were assessed in a subset of 59 participants with PBMC samples, contributing 172 samples. At day 0, the frequency of IFN-γ secreting cells to overlapping SARS-CoV-2 spike peptide was significantly higher in asymptomatic patients compared to severe patients (GMs: 117 [95% CI 71–194] vs 32 [95% CI 8–76]; p=0.0366) (Figure 1a, Table S3). For the IFN-γ secreting cells specific to overlapping M peptides, severe patients exhibited significantly higher levels at day 7 than mild/moderate patients (GMs: 90 [95% CI 37–217] vs 19 [95% CI 10–39]; p= 0.0180; Figure 1b, Table S3). We did not observe any significant differences in the frequencies of IFN-γ secreting cell responses to NP, NSPs, and ORFs overlapping peptides (Figure 1c–e, Table S3). For all five SARS-CoV-2 peptides we observed temporal differences in IFN-γ secreting cells within each COVID-19 severity group (Figure 1, Table S3). Spike peptide: In the asymptomatic group, IFN-γ secreting cells were significantly elevated on day 7 than on day 14 (GMs: 161 [95% CI 86–301] vs 92 [95% CI 44–189]; p=0.0039) and day 28 (102 [95% CI 49–209]; p=0.0042). For the mild/moderate group, levels significantly increased from 86 (95% CI 53–138) on day 7 to 142 (95% CI 78–257) on day 14 (p=0.0403). In the severe group, significantly higher levels were observed on day 7 (70 [95% CI 29–165]; p=0.0381), day 14 (117 [95% CI 52–260]; p=0.0058), and day 28 (108 [95% CI 33–348]; p=0.0037) relative to day 0 (23 95% CI 7–76).

**Figure 1.**
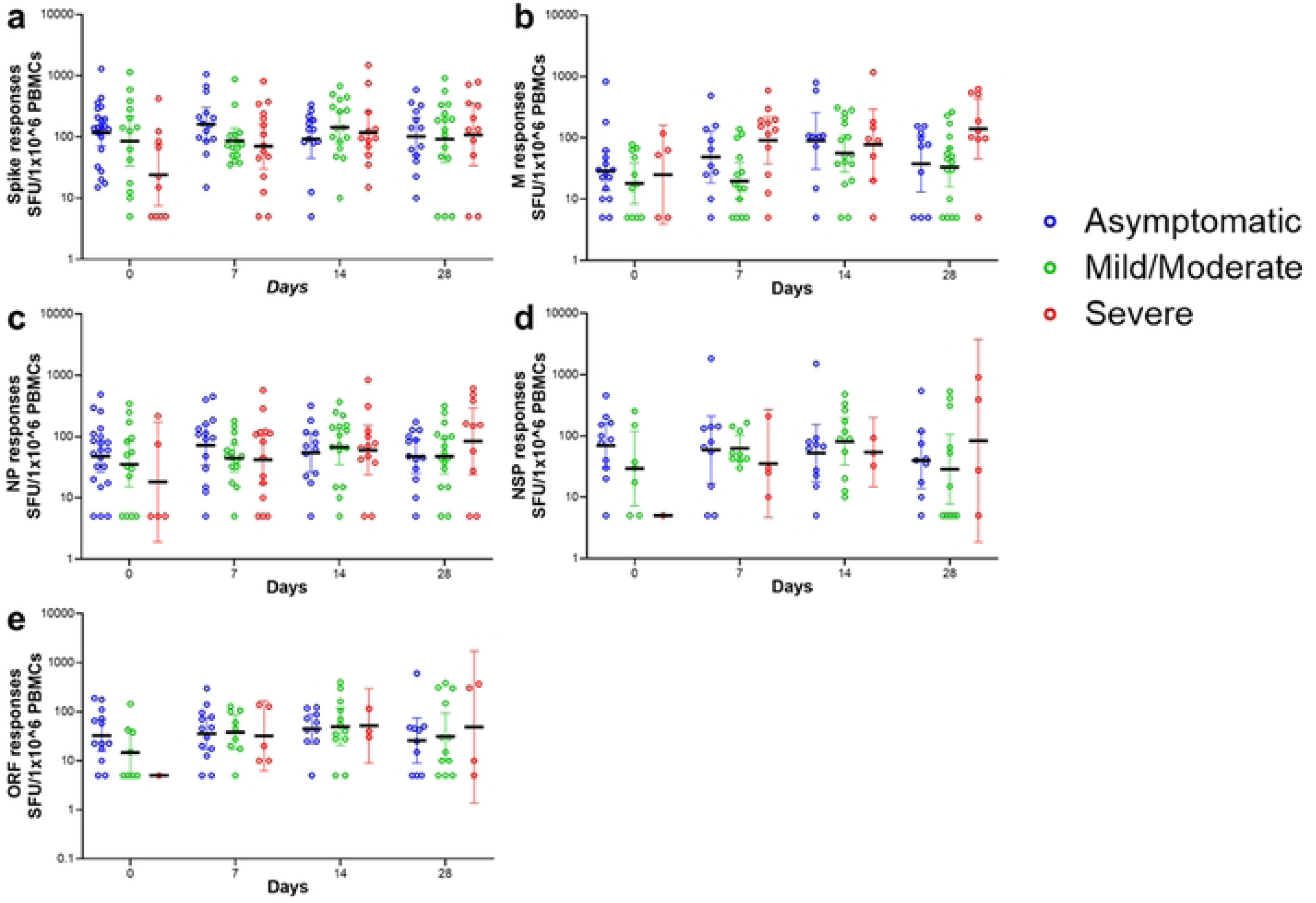
Induction of a higher IFN-gamma cellular response to Spike protein on day of diagnosis is associated with asymptomatic infection. Frequencies of ex-vivo IFN-γ secreting cells against SARS-CoV-2 peptide pools spanning (**a)** spike protein, (**b)** M protein, **c** NP protein, (**d)** NSP proteins and (**e)** ORF proteins. Bars represent geometric mean and 95% CI. Linear mixed effects model with Tukey’s multiple comparisons, was used, * P < 0.05. Number of samples analysed for: spike responses = 171, M responses = 136, NP responses = 162, NSP responses = 100 and ORF responses = 90.

M Peptide: In the asymptomatic patients, IFN-γ secreting cells were significantly higher on day 14 (89 [95% CI 30–258]) compared to day 0 (GMs: 28 [95% CI 13–59]; p=0.0364) and day 7 (GMs: 48 [95% CI 18–129]; p=0.0297). In the mild/moderate group, day 14 levels were significantly higher compared to day 0 (p=0.0039), day 7 (p<0.0001), and day 28 (p=0.002). Additionally, day 28 levels were significantly higher than day 7 (p=0.0049). No significant differences were observed between timepoints in the severe group.

NP Peptide: Among the asymptomatic group, a significant decline was observed from day 7 (71 [95% CI 33–153]) to day 28 (47 [95% CI 24–88]; p=0.0035). In the mild/moderate group, IFN-γ secreting cell levels were significantly higher on day 14 (GMs: 67 [95% CI 34– 130]) compared to day 0 (GMs: 35 [95% CI 14–83]; p=0.0037), day 7 (GMs: 44 [95% CI 26–73]; p=0.0178), and day 28 (GMs: 47 [95% CI 24–91]; p=0.0003). No significant differences were observed between timepoints in the severe group. For NSP peptide: a significant decline in IFN-γ secreting cells was observed within the mild/moderate group from 80 (95% CI 33– 194) at day 14 to 28 (95% CI 7–106) at day 28 (p=0.0316). For ORF peptide: a significant increase in IFN-γ secreting cells was observed in the mild/moderate group between day 0 (14 [95% CI 4–43]) and day 7 (GMs: 38 [95% CI 16–85]; p=0.0242).

We also assessed whether there were differences in IFN-γ secreting cell responses by location (urban Nairobi and rural Kilifi) among asymptomatic and mild patients. Relative to Nairobi, significantly higher levels of IFN-γ secreting cell responses to the SARS-CoV-2 spike peptide (218 [95% CI 125–381] vs 56 [95% CI 28–110]; p=0.0057; Figure 2a) and NP peptide (94 [95% CI 51–169] vs 19 [95% CI 8–48; p=0.0171; Figure 2c) were observed among the asymptomatic patients in Kilifi on day 0. There were no significant differences between Kilifi and Nairobi in the IFN-γ secreting cell responses to overlapping peptides for the M protein for the asymptomatic patients (Figure 2b), nor for the spike, or NP, and or M peptides among mild patients (Figure 2d–f). Data for severe patients are shown for Nairobi participants only (Figure 2g–i), as we were unable to recruit severe patients in Kilifi.

**Figure 2.**
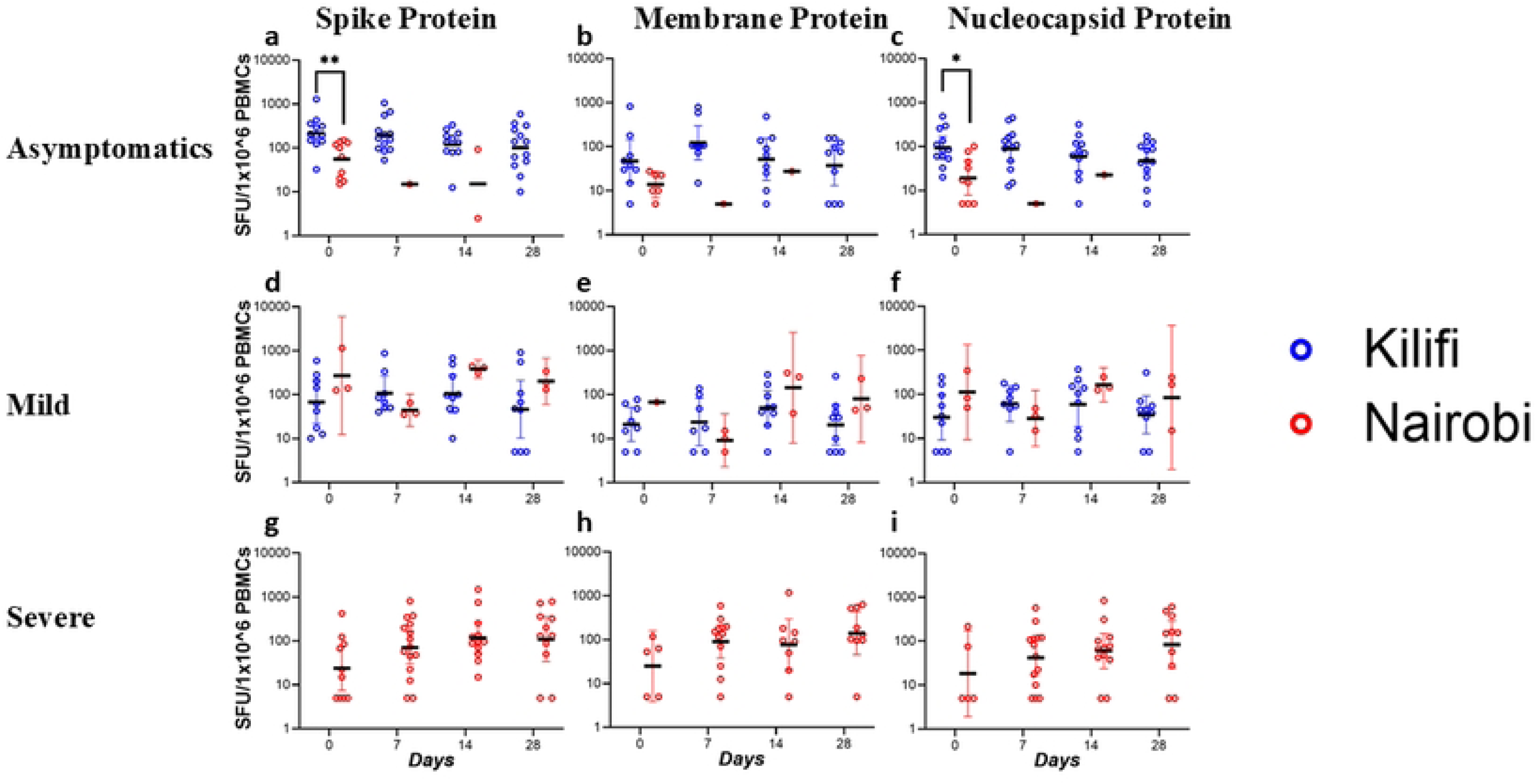
Induction of a higher IFN-γ cellular response on the day of diagnosis is associated with Kilifi participants. Comparison of IFN-γ cellular responses between Kilifi and Nairobi COVID-19 patients with: asymptomatic disease for (**a)** Spike protein, (**b)** M protein, (**c)** NP protein; Mild disease for (**d)** Spike protein, € M protein, (**f)** NP protein; and Severe disease for (**g)** Spike protein, (**h)** M protein, (**i)** NP protein. Kilifi didn’t have severe patients. Bars represent geometric mean and 95% CI. Kruskal–Wallis one-way ANOVA, with Dunn’s multiple comparisons test, was performed. * P < 0.05, **P < 0.01.

### Kinetics of cytokine and chemokine responses across COVID-19 severity groups and geographical location

Asymptomatic patients consistently showed elevated levels of SDF-1α at all time-points, but lower levels of all the other cytokines and chemokines measured and no detectable levels of IL-9 (Figure 3a). Similarly, for mild/moderate patients, high levels of SDF-1α (Figure 3b), were observed at all timepoints whereas other cytokines and chemokines were secreted at low to intermediate levels. High cytokine and chemokines levels were seen among the severe patients with IL-Iβ, IL-6, IL-2, IFN-γ, GM-CSF, IFN-α, IL-7 and GRO-α decreasing over time, while MIP-1α, MIP-Iβ, MCP-1, and SDF-1α increased with time. Levels for IL1-ra, and IL-9 were similar at all the time points (Figure 3c).

**Figure 3.**
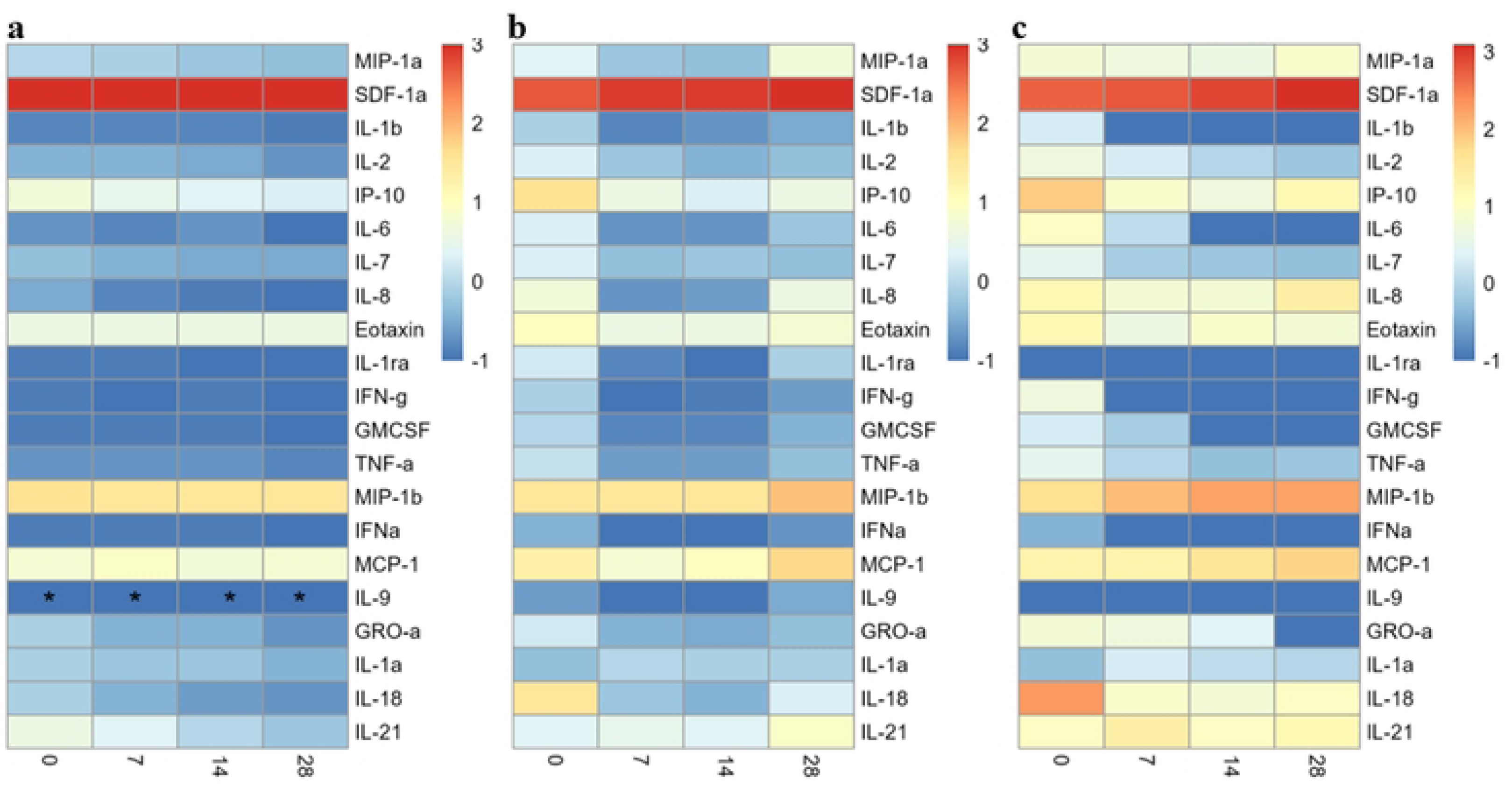
Kinetics of Cytokine and Chemokine Concentrations in COVID-19 Patients. Mean of log10-transformed cytokine/chemokine concentrations plotted over time for **(a)** Asymptomatic participants, (**b)** Mild/Moderate participants, and (**c)** Severe participants. * - Levels for all participants were below detectable levels.

We compared asymptomatic against mild/moderate and severe participants to evaluate differences among the clinical phenotypes. Mild/moderate participants had significantly higher levels of IL-18, IL-8, IL-1ra, IL-6, GM-CSF, IP-10, MCP-1, TNF-α, MIP-1α, IFN-γ, IL-2, IL-7, IL-1β, IL-9, Eotaxin and IFN-α than asymptomatic patients. The largest effect sizes were observed with IL-18 (1.107, p<0.0001), IL-8 (1.105, p<0.0001) and IL-1ra, (0.672, p=0.013). On the contrary, levels for SDF-1α were significantly reduced among the mild/moderate patients relative to the asymptomatic (effect size -0.182, p<0.0001) (Figure 4). Severe participants had significantly higher levels for cytokines (IL-8, IL-18, IL-1ra, IL-6, IP-10, MIP-1α, TNF-α, IL-9, IFN-γ, GM-CSF, IL-7, IL-1β, MCP-1, IL-2, GRO-α, Eotaxin and IFN-α) compared to asymptomatic cases, except for IL-1a, IL-21 and MIP-Iβ which had similar levels, and SDF-1α (effect size -0.195, p<0.0001), which was significantly reduced. The largest effect size was observed in IL-8 (1.754, p<0.0001), IL-18 (1.666, p<0.0001) and IL-1ra (1.197, p<0.0001) (Figure 4).

**Figure 4.**
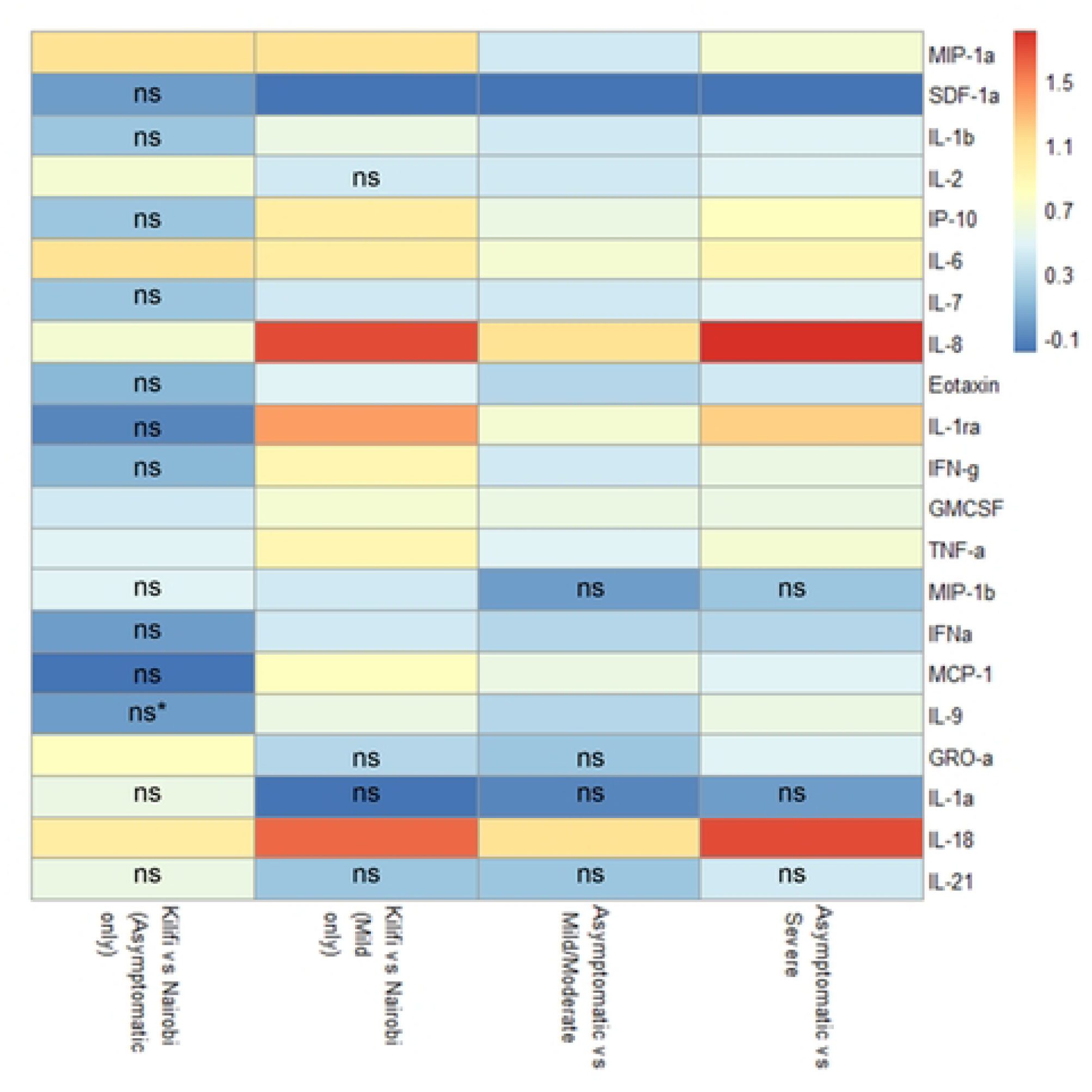
Higher Cytokine and Chemokine concentrations in Asymptomatic and Mild Patients from Nairobi Compared to Kilifi, and in Mild/Moderate and Severe Patients Compared to Asymptomatic Individuals. ns – not-significant, *p* value >0.05, empty means it was significant at p<0.05 - <0.0001. * - Levels for all participants were below detectable levels.

For asymptomatic individuals, the cytokines and chemokine levels for IL-6, MIP-1α, IL-18, GRO-α, IL-2, IL-8, TNF-α and GM-CSF were significantly higher among Nairobi than Kilifi patients. Comparisons for IL-6 (1.139, p<0.0001), MIP-1a (1.093, p=0.004) and IL-18 (1.025, p=0.002) had the largest effect sizes (Figure 4). For mild patients, all cytokines (IL-8, IL-18, IL-1ra, MIP-1α, IL-6, IP-10, IFN-γ, TNF-α, MCP-1, GM-CSF, IL-9, IL-1β, Eotaxin, IL-7, MIP-Iβ, and IFN-α) were significantly higher among Nairobi patients in comparison to Kilifi patients except for GRO-α, IL-1α, IL-2 and IL-21 which were similar. Notably, the largest effect size for these comparisons was observed for IL-8 (1.69, p<0.0001), IL-18 (1.634, p<0.0001) and IL-1ra (1.355, p<0.001). On the other hand, SDF-1α (effect size - 0.161, p = 0.017) was significantly lower in Nairobi than in Kilifi patients.

#### Principal component analysis

The concentrations of the majority of the 21 cytokines and chemokines were positively correlated with each other. However, the levels of SDF-1α were negatively correlated with those of IL-2, IP-10, IL-7, IFN-γ, GM-CSF, and IL-18 (Figure S1). We retained the first three principal components from a principal component analysis, accounting for 62% of the total variability in the 21 cytokines and chemokines (Figure S2a). IL-9, MIP-1α, TNF-α, MCP-1, MIP-Iβ, IL1-ra, IL-6, IL-Iβ, GRO-α and IL-8 were the most strongly associated with the first PC. The second PC was most strongly associated with IP-10, IFN-γ, GM-CSF, IL-2, IL-18, IL-7, IFN-α, Eotaxin and SDF-1α. The third PC was associated with IL-1 α and IL-21 (Figure S3). There was no clear separation among the mild/moderate and severe groups. However, data points for the asymptomatic participants clustered together demonstrating a much-reduced diversity of cytokine and chemokine levels than the other groups (Figure 5), and median PC scores did not change over the first month (Figure S2b).

**Figure 5.**
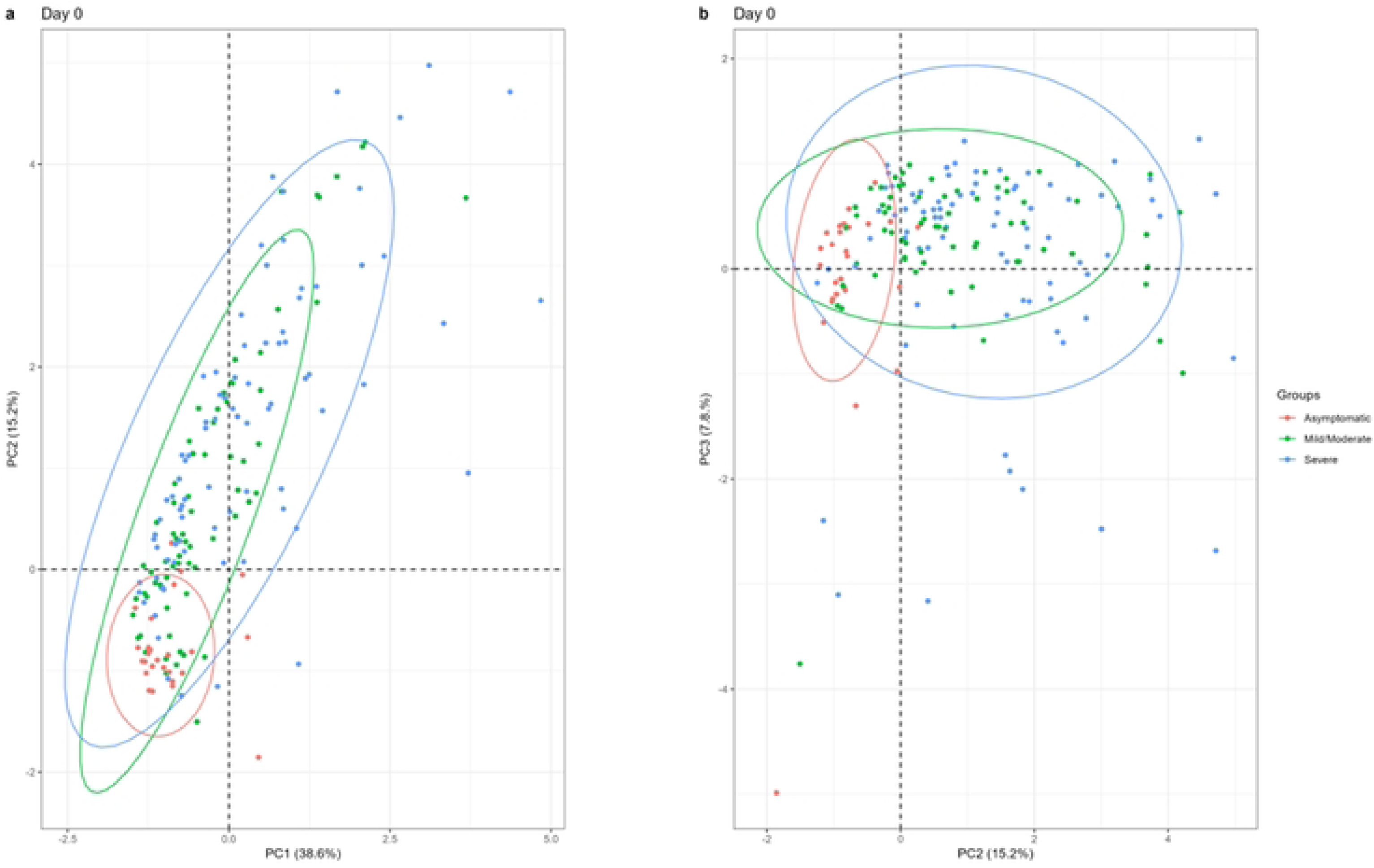
Scatter plot by Severity at baseline. **(a)** PC1 vs PC2; (**b)** PC2 vs PC3

Among the symptomatic groups (mild/moderate/severe), a steady decline of all the PCs over one month was observed, with a steep decline occurring between day 0 and day 7 (Figure S2c). There was no apparent difference of the cytokine and chemokine responses between asymptomatic participants from Kilifi and Nairobi (Figure S4a). For mild cases, Kilifi patients clustered together, although there was a slight overlap with Nairobi participants (Figure S4b). Using biplots, there was no apparent difference between asymptomatic and mild Kilifi patients (Figure S4c). For Nairobi, clinical phenotypes were asymptomatic, mild, moderate, and severe. We combined the mild and moderate, and observed no clear difference between the mild/moderate and severe groups, while the few asymptomatic participants seemed to cluster together (Figure S4d).

#### Discussion and conclusion

In this study, we measured associations between the acute IFN-γ cellular, and 22 cytokine and chemokine, immune response to SARS-CoV-2 (within a month of diagnosis) with COVID-19 severity and possible modulation by differential environmental exposures in urban and rural areas in Kenya.

As had been seen in earlier reports from Singapore, Netherlands, and Italy, which associated increased IFN-γ secreting cells measured by ELISpot assay in the early phase of infection with milder disease (38–40), we found that higher frequencies of an early IFN-γ cellular response to SARS-CoV-2 spike overlapping peptides was associated with asymptomatic, compared to severe SARS-CoV-2 infection outcomes. This observation would suggest either: 1) that the IFN-γ cellular response contribute to protection against disease progression, or 2) that developing severe COVID-19 depresses this response. The latter interpretation may be supported by previous reports from China and USA reporting correlations of an early induction of increased basal T-cell specific responses (CD4^+^ and CD8^+^ T) in COVID-19 patients with mild disease, which was suppressed among their severely sick counterparts (41,42).

Whilst the concentrations of the pro-inflammatory cytokines and chemokines IL-6, IL-Iβ, GRO-α, IFN-γ, GM-CSF, IFN-α, IL-7 and IL-2 in severe patients declined to basal levels at day 28 from day of diagnosis, we found that the concentrations for the chemokines MIP-1α, MIP-Iβ, MCP-1, and SDF-1α were increasing with time. This is to be expected as cytokines and chemokines play different roles and at different times of the immune response to infection. MIP-1α, MCP-1 and MIP-Iβ are chemoattractant and play key roles in the recruitments of leukocytes such as monocytes, T-cells, and neutrophil to the sites of infection (43,44). Similarly increasing kinetics for MIP-1α and MCP-1 in severe and fatal patients were reported in Norway and China, respectively (45,46). Notably, we found that higher concentrations of SDF-1α were associated with asymptomatic individuals, hinting at a potential protective role from severe disease progression. SDF-1α (CXCL12), is a chemokine involved in the recruitment of T-cells (47), CD34+ hematopoietic stem/progenitor cells (48), lymphocytes and monocytes (49) to the site of infection further enhancing inflammation. In contrast, studies from China and Bulgaria did not observe significant differences in SDF-1α levels among asymptomatic, mild, moderate, severe or fatal cases (18,46,50). However, others have also implicated SDF-1α in disease severity based on genetic association studies in a single-centre study (51), although this finding was not corroborated in multi-centre genome-wide association studies (52). SDF-1α may aid in the timely recruitment of T and other cells (47), to the sites of infection, enhancing viral elimination, reduction in inflammation, and promotion of recovery.

We demonstrate that elevated levels of eighteen cytokines and chemokines were associated with severe COVID-19, in agreement with previous reports linking them to severe lung injury and ARDS (17–22,53). However, the associations with IL-8, IL-18 and IL-1ra, had the strongest effect sizes in the current study. IL-8, has been implicated in the activation and recruitment of neutrophils to the site of infection, thereby promoting inflammation (54). IL-18 amplifies the immune response by inducing the production of IFN-γ by T-cells and natural killer cells (55). Thus, IL-8 and IL-18 could amplify the excessive inflammation that characterises severe COVID-19. On the contrary, IL1-ra is known to suppress the production of proinflammatory cytokines such as IL-1 and TNF-α (56), and probably helps mitigate the effects of excessive inflammation, thus reducing tissue damage and associated mortality.

In parallel to the reduced rates of severe COVID-19 and associated deaths in SSA relative to North America and Europe, we and others have suggested higher rates of severe COVID-19 in cities compared to rural areas in SSA (28). Whilst this difference could be explained by higher levels of SARS-CoV-2 transmission in busy metropolitan cities, or by more complete reporting of cases (28,57,58), we wondered whether there were also plausible biological explanations. We compared inflammatory cytokine and chemokine levels, and found that the asymptomatic patients from Nairobi had higher levels of 8 cytokines and chemokines than their asymptomatic counterparts from Kilifi, with IL-6, IL-18 and MIP-1α, having the strongest associations. Similarly, levels of 16 cytokines and chemokines were higher among mild-Nairobi, than Kilifi, COVID-19 patients with IL-8, IL-18 and IL-1ra being the most differentially secreted. Moreover, the basal frequencies of IFN-γ secreting cells in asymptomatic patients from Nairobi, relative to those of their Kilifi counterparts, were reduced. Collectively, these findings would suggest that the immune response to SARS-CoV-2 is less inflammatory among residents of rural areas (59).

Our study was faced with a few limitations. Firstly, our data are incomplete for some of the patients due to missed follow-ups, or due to unavailability of adequate numbers of PBMCs to quantify IFN-γ cellular responses to the full spectrum of the peptide pools corresponding to all the different SARS-CoV-2 proteins. However, our analyses are assumed valid under the missing at random mechanism given the likelihood approach (60). Secondly, we had difficulties recruiting asymptomatic patients in Nairobi and were unable to recruit severe cases in Kilifi and thus the sample size is relatively small.

In conclusion, although severe disease was rare in Kenya, the inflammatory cytokine profile in patients with severe COVID is similar to that of North American and European severe COVID-19 patients. However, just like the early IFN-γ secreting cellular response, increased levels of the chemokine, SDF-1α, were associated with asymptomatic SARS-CoV-2 infection, suggesting a potential role of these responses in protection against disease progression. Finally, the differential cytokine and chemokine, and IFN-γ cellular responses between urban Nairobi and rural Kilifi patients suggest a plausible biological explanation for the increased frequency of severe COVID-19 in African SSA cities relative to rural areas. Together, these findings provide insights into potentially COVID-19 protective immune responses, and their modulation by differential environmental exposures. Nonetheless, additional studies are required to extend and replicate these important findings as they could inform future control of COVID-19 and empower the control of similar pandemics.

## Data Availability

All data files are available from the Harvard Dataverse at the KWTRP Research Data Repository https://doi.org/10.7910/DVN/XMTCTW

## Acknowledgments

We thank the field workers, laboratory staff and healthcare workers involved in the longitudinal blood sampling. We appreciate all the study participants.

## Supporting information

**Figure S1. Correlation matrix of 21 cytokines across all time points from 400 patients.** Pearson’s correlation coefficients are visualised, with red indicating positive correlations, blue negative correlations, and white representing no correlation.

**Figure S2. Principal component analysis of the cytokines. a)** Scree-plot showing that the first 3 principal components explained 62% of variability in the cytokines data for all participants; **(b)** Line plots for the first 3 principal component illustrating trends over time for asymptomatic participants; and **(c)** Line plots for the first 3 principal component illustrating trends over time for symptomatic (mild, moderate and severe) participants.

**Figure S3. Cytokines loading on the first three principal components (PC1–PC3).** The color scale indicates the loading value, with red indicating a higher positive loading, blue a higher negative loading and light-yellow minimal loading.

**Figure S4. Scatter plots showing cytokine data for participants grouped by location and clinical phenotype**. **(a)** Asymptomatic participants, **(b)** mild cases, **(c)** participants from Kilifi, and **(d)** participants from Nairobi. Each point represents an individual’s cytokine measurement, allowing for a visual assessment of cytokine variability across different groups based on location and clinical presentation.

**Table S1. Peptides Sequences.** Each entry represents a distinct peptide along with its associated properties. The pooling strategy for the 10 peptide pools used is shown below.

**Table S2. Sample size for each peptide.** Number of participants sampled for each peptide across different clinical phenotypes (asymptomatic, mild/moderate, severe) and time points (Day 0, 7, 14, 28).

**Table S3. Associations of SARS-CoV-2 Peptides (Spike, M, NP, NSP, and ORF) with asymptomatic, mild/moderate, and severe clinical phenotypes over time. Tukey’s** multiple comparisons test was used to assess differences within and between clinical phenotypes across different time points. Bold p values are significant, p<0.05.

## Notes

### Competing Interest Statement

The authors have declared no competing interest.

### Clinical Trial

"N/A''

### Funding Statement

Yes

### Author Declarations

Kenya Medical Research Institute’s Scientific and Ethics Review Unit and the Aga Khan University, Nairobi, Institutional Ethics Review Committee .

